# Validation of Saliva and Self-Administered Nasal Swabs for COVID-19 Testing

**DOI:** 10.1101/2020.08.13.20173807

**Authors:** Alvin Kuo Jing Teo, Yukti Choudhury, Iain Beehuat Tan, Chae Yin Cher, Shi Hao Chew, Zi Yi Wan, Lionel Tim Ee Cheng, Lynette Lin Ean Oon, Min Han Tan, Kian Sing Chan, Li Yang Hsu

**Author notes:** Corresponding author: Li Yang Hsu, Address: Saw Swee Hock School of Public Health, National University of Singapore, #10-01, 12 Science Drive 2, 117549 Singapore.

## Abstract

**Background:** Active cases of COVID-19 has primarily been diagnosed via RT-PCR of nasopharyngeal (NP) swabs. Saliva and self-administered nasal (SN) swabs can be collected safely without trained staff.

We aimed to test the sensitivity of “naso-oropharyngeal” saliva and SN swabs compared to NP swabs in a large cohort of migrant workers in Singapore.

**Methods:** We recruited 200 male adult subjects: 45 with acute respiratory infection, 104 asymptomatic close contacts, and 51 confirmed COVID-19 cases. Each subject underwent NP swab, SN swab and saliva collection for RT-PCR testing at 1 to 3 timepoints. We additionally used a direct-from-sample amplicon-based next-generation sequencing (NGS) workflow to establish phylogeny.

**Results:** Of 200 subjects, 91 and 46 completed second and third rounds of testing, respectively. Of 337 sets of tests, there were 150 (44.5%) positive NP swabs, 127 (37.7%) positive SN swabs, and 209 (62.0%) positive saliva.

Test concordance between different sample sites was good, with a kappa statistic of 0.616 for NP and SN swabs, and 0.537 for NP and saliva. In confirmed symptomatic COVID-19 subjects, the likelihood of a positive test from any sample fell beyond 14 days of symptom onset.

NGS was conducted on 18 SN and saliva samples, with phylogenetic analyses demonstrating lineages for all samples tested were Clade O (GISAID nomenclature) and lineage B.6 (PANGOLIN nomenclature).

**Conclusion:** This study supports saliva as a sensitive and less intrusive sample for COVID-19 diagnosis and further delineates the role of oropharyngeal secretions in increasing the sensitivity of testing. However, SN swabs were inferior as an alternate sample type. Our study also provides evidence that a straightforward next-generation sequencing workflow can provide direct-from-sample phylogenetic analysis for public health decision-making.

## Introduction

The severe acute respiratory syndrome coronavirus 2 (SARS-CoV-2) emerged from Wuhan, China in November 2019 [1], and has since caused a global pandemic, with over 13.4 million confirmed COVID-19 cases and 581,000 deaths as of 15^th^ July 2020 [2]. Singapore has not been spared, with over 46,000 cases and 27 deaths since the first case was reported on 23^rd^ January 2020, the majority of cases being migrant workers living in crowded dormitories [3].

The principal method for diagnosing active cases of coronavirus disease 2019 (COVID-19) has been through the detection of viral genetic material via reverse transcription-polymerase chain reaction (RT-PCR). In Singapore and several other countries, nasopharyngeal (NP) swabs are the principal means for collecting specimens for testing [4,5]. However, NP swabs could cause discomfort and require trained healthcare staff to perform. In situations where mass or community screening is required, the resources required to organize NP swabbing can be considerable, such as the drive-through screening centres pioneered by South Korea [6].

Saliva and self-administered nasal (SN) swabs are, in many ways, ideal specimens for COVID-19 screening. Both can be collected safely without the need for trained staff. The utility of saliva for COVID-19 testing has been tested in multiple territories and countries, including Hong Kong [7–9], China [10], Italy [11], Australia [12], North America [13,14], and Japan [15].

The majority of current published studies involve relatively small numbers of subjects with heterogeneous results, and a meta-analysis suggests that saliva is at best slightly less sensitive or similar to other specimens, including NP swabs [16]. One caveat relates to how saliva is collected—saliva is a complex bio-mixture which can consist of salivary gland secretion, gingival crevicular fluid, sputum and/or mucosal transudate, in varying proportions depending on collection method. Some studies tested only secretions from the mouth [11,12], others explicitly tested “posterior oropharyngeal” or “deep throat” saliva with secretions from the oropharynx [7–10], while the rest were unspecified [13–16].

We aimed to test the sensitivity of “naso-oropharyngeal” saliva and SN swabs compared to NP swabs in a large cohort of migrant workers in Singapore using RT-PCR testing. We additionally used a direct-from-sample amplicon-based next-generation sequencing (NGS) workflow to establish phylogeny for tested samples.

## Methods

### Study Population

Subjects were recruited from two sites – a large 5,400-bed purpose-built dormitory where migrant workers were housed in large rooms holding between 7 and 20 workers, and a community care facility (CCF) where migrant workers diagnosed with COVID-19 via prior RT-PCR testing but not requiring acute hospital care were sent for monitoring and isolation. All subjects at the CCF are prior confirmed cases, while subjects from the dormitory comprised of two groups—1) migrant workers presenting with symptoms of acute respiratory tract infection (ARI); and 2) asymptomatic roommates of newly diagnosed COVID-19 cases.

### Sample Collection

Migrant workers from the purpose-built dormitory presenting with ARI were assessed at the medical post, where the decision for NP swab for diagnosis of COVID-19 was made by the physician-in-charge. After their routine nasopharyngeal swabs were obtained by a trained healthcare worker, these workers were immediately approached for study participation, and consent taken where agreeable. Demonstration videos on the performance of SN swab and naso-oropharyngeal saliva collection in the major native languages of the migrant workers (English *video link: https://youtu.be/4jGrJUbjBBs)* were shown, following which these samples were collected under the supervision of a trained researcher. For other groups of consenting subjects from community care facility (CCF) previously diagnosed with COVID-19, the NP swab was performed by a trained researcher following collection of the SN swab and naso-oropharyngeal saliva in the same sitting.

We planned for each subject to be tested up to three times at 2-3-day intervals where possible, in order to compare the sensitivity of different samples across time. Subjects from the purpose-built dormitory who tested negative across all three samples during the first round of testing were not retested. Subjects from the CCF were not retested if all samples from the initial two rounds were negative.

NP swabs from subjects with ARI were sent dry in cooler boxes to the Singapore General Hospital (SGH) molecular laboratory as part of routine clinical testing and processed within the same day. NP swabs and SN swabs from other subjects were sent in 3 mis of viral transport medium, while between 1-2 mis of oropharyngeal saliva was collected in a container with 2 mls of viral RNA stabilization fluid (SAFER™-Sample Stabilization Fluid, Lucence, Singapore) before transfer to Lucence Laboratory with processing occurring within the same day. Both service laboratories are College of American Pathologists (CAP) accredited.

### Laboratory Testing

RT-PCR at SGH was performed using the cobas®SARS-CoV-2 (Roche Molecular Systems, Branchburg, NJ) on an automated cobas®6800 system, with results called according to the manufacturer’s specifications.

NP and saliva samples sent to Lucence Laboratory underwent RNA extraction (200 μl of the sample) (GeneAid Biotech Ltd) and were tested with a laboratory-developed RT-PCR test (CDC-LDT) based on primers published by the Division of Viral Diseases, National Center for Immunization and Respiratory Diseases, Centers for Disease Control and Prevention, Atlanta, GA, USA [17], while saliva and SN swabs were additionally tested using the Fortitude 2.1 kit (MiRXES, Singapore). The analytical limit of detection of the CDC-LDT was determined to be 25 copies per reaction based on a synthetic SARS-CoV-2 genome (Twist Bioscience). Saliva was pre-processed with the addition of dithiothreitol (DTT), vortexing and incubation at room temperature for 15 minutes.

SARS-CoV-2 amplicon-based next-generation sequencing (NGS) was done for 18 available samples collected in stabilization fluid from known COVID-19 cases, using 330 primer pairs to generate amplicons (size range 130-178 bp) covering the entire virus genome (except the first 25 bases and 30 bases upstream of the final polyA tail) to establish a direct-from-sample workflow. To rule out potential non-specific amplification of other viruses related in sequence, all amplicons were verified to have limited similarity to sarbecoviruses, outside of SARS-related coronaviruses (assumed to not be in present circulation). Ten of 18 samples were paired sets of saliva and SN swab samples from 5 individuals. The remaining 8 comprised 1 SN swab and 7 saliva samples from individual patients. For samples with complete viral genomes (100% coverage ≥ 1X coverage), phylogenetic analysis was performed to identify lineages based on sequence variants.

### Statistical Methods

Intercooled Stata 13.1 (StataCorp, College Station, TX, USA) was used for all statistical calculations. The kappa statistic was used to measure observer agreement between sample sites.

### Ethics

This study was approved by the Director of Medical Services, Ministry of Health under Singapore’s Infectious Disease Act [18].

### Funding Source

This study was funded by a grant from the Changi Airport Group. Fortitude 2.1 kits were provided by the Diagnostics Development Hub (DxD Hub), A*ccelerate, Agency for Science, Technology and Research, Singapore. The study sponsor had no role in the design, implementation, analysis or write-up of the study.

## Results

Study recruitment was completed between 2^nd^ and 26^th^ June 2020. There were 45 subjects with ARI and 104 asymptomatic close contacts recruited from the purpose-built dormitory, while 51 subjects with confirmed COVID-19 (8 asymptomatic at the time of diagnosis) were recruited at the CCF.

Subject characteristics are displayed in Table 1. Of 200 subjects, 91 and 46 completed second and third rounds of testing, respectively. Because COVID-19-positive migrant workers were rapidly transferred out of dormitories to CCFs, all but one subject from the dormitory site did not complete the planned testing. The median time period between the date of diagnosis and the first round of testing in asymptomatic subjects was 0 days (range: 0-6 days) while the median time period between symptom onset and the first round of testing was 5.5 days (range: 0-28 days).

**Table 1.**
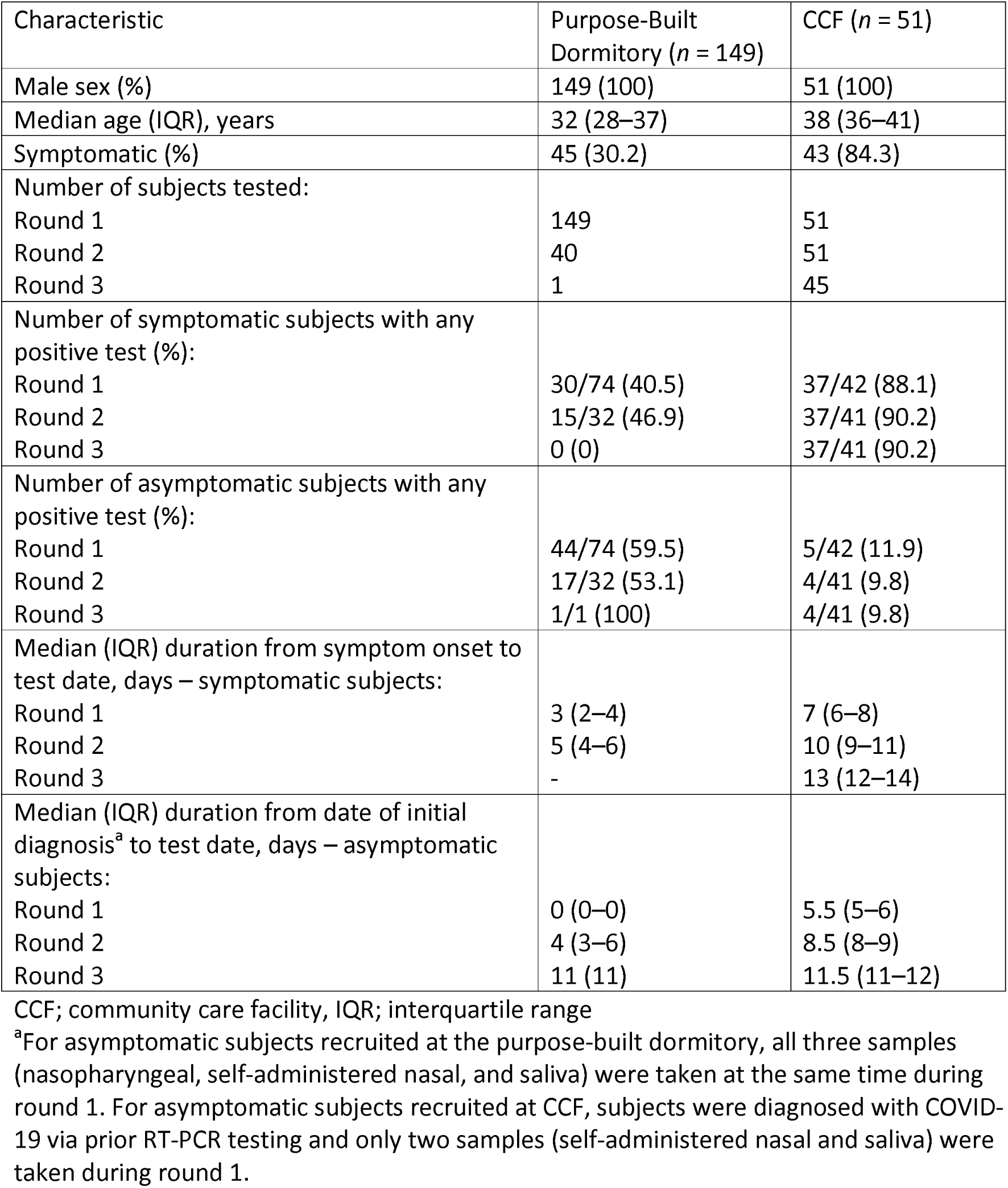
Characteristics of recruited subjects.

| Characteristic | Purpose-Built<br>Dormitory (*n* = 149) | CCF (*n* = 51) |
| --- | --- | --- |
| Male sex (%) | 149 (100) | 51 (100) |
| Median age (IQR), years | 32 (28–37) | 38 (36–41) |
| Symptomatic (%) | 45 (30.2) | 43 (84.3) |
| Number of subjects tested: |  |  |
| Round 1 | 149 | 51 |
| Round 2 | 40 | 51 |
| Round 3 | 1 | 45 |
| Number of symptomatic subjects with any<br>positive test (%): |  |  |
| Round 1 | 30/74 (40.5) | 37/42 (88.1) |
| Round 2 | 15/32 (46.9) | 37/41 (90.2) |
| Round 3 | 0 (0) | 37/41 (90.2) |
| Number of asymptomatic subjects with any<br>positive test (%): |  |  |
| Round 1 | 44/74 (59.5) | 5/42 (11.9) |
| Round 2 | 17/32 (53.1) | 4/41 (9.8) |
| Round 3 | 1/1 (100) | 4/41 (9.8) |
| Median (IQR) duration from symptom onset to<br>test date, days – symptomatic subjects: |  |  |
| Round 1 | 3 (2–4) | 7 (6–8) |
| Round 2 | 5 (4–6) | 10 (9–11) |
| Round 3 | - | 13 (12–14) |
| Median (IQR) duration from date of initial<br>diagnosis<sup>a</sup> to test date, days – asymptomatic<br>subjects: |  |  |
| Round 1 | 0 (0–0) | 5.5 (5–6) |
| Round 2 | 4 (3–6) | 8.5 (8–9) |
| Round 3 | 11 (11) | 11.5 (11–12) |

CCF; community care facility, IQR; interquartile range
<sup>a</sup>For asymptomatic subjects recruited at the purpose-built dormitory, all three samples (nasopharyngeal, self-administered nasal, and saliva) were taken at the same time during round 1. For asymptomatic subjects recruited at CCF, subjects were diagnosed with COVID-19 via prior RT-PCR testing and only two samples (self-administered nasal and saliva) were taken during round 1.

The results of RT-PCR testing are shown in Table 2. Of 337 sets of tests, there were 150 (44.5%) positive NP swabs tested via cobas®SARS-CoV-2 or CDC-LDT, 127 (37.7%) positive SN swabs tested via CDC-LDT, 119 (35.3%) positive SN swabs tested via Fortitude 2.1, 209 (62.0%) positive saliva tested via CDC-LDT, and 167 (49.6%) positive saliva tested via Fortitude 2.1. Ct values were generally lower during the earlier period of infection across all sample types, in particular for symptomatic infections where the onset of illness could be better estimated (Figure 1).

**Table 2.** Results of RT-PCR testing on samples.

|  | Tests on symptomatic individuals (*n* = 188) | Tests on asymptomatic individuals (*n* = 149) |
| --- | --- | --- |
| Positive results, *n* (%): |  |  |
| NP swab - CDC-LDT | 123 (65.4) | 27 (18.1) |
| SN swab - CDC-LDT | 96 (51.1) | 31 (20.8) |
| SN swab - Fortitude 2.1 | 98 (52.1) | 21 (14.1) |
| Saliva – CDC-LDT | 145 (77.1) | 64 (43.0) |
| Saliva – Fortitude 2.1 | 123 (65.4) | 44 (29.5) |
| Concordance of results among NP swabs (CDC-LDT), SN swabs (CDC-LDT), and saliva (CDC-LDT), *n* (%): |  |  |
| NP swab only positive | 6 (3.2) | 1 (0.7) |
| Saliva only positive | 24 (12.8) | 35 (23.5) |
| SN swab only positive | 2 (1.1) | 8 (5.4) |
| NP swab and saliva positive | 29 (15.4) | 8 (5.4) |
| NP and SN swab positive | 2 (1.1) | 2 (1.3) |
| Saliva and SN swab positive | 6 (3.2) | 5 (3.4) |
| All three swabs positive in-tandem | 86 (45.7) | 16 (10.7) |
| Concordance of results among NP swabs (CDC-LDT), SN swabs (Fortitude 2.1), and saliva (CDC-LDT), *n* (%): |  |  |
| NP swab only positive | 5 (2.7) | 1 (0.7) |
| Saliva only positive | 25 (13.3) | 39 (26.2) |
| SN swab only positive | 1 (0.5) | 4 (2.7) |
| NP swab and saliva positive | 26 (13.8) | 10 (6.7) |
| NP and SN swab positive | 3 (1.6) | 2 (1.3) |
| Saliva and SN swab positive | 5 (2.7) | 1 (0.7) |
| All three swabs positive in-tandem | 89 (47.3) | 14 (9.4) |
| Concordance of results among NP swabs (CDC-LDT), SN swabs (CDC-LDT), and saliva (Fortitude 2.1), *n* (%): |  |  |
| NP swab only positive | 12 (6.4) | 4 (2.7) |
| Saliva only positive | 14 (7.5) | 20 (13.4) |
| SN swab only positive | 4 (2.1) | 9 (6.0) |
| NP swab and saliva positive | 22 (11.7) | 5 (3.4) |
| NP and SN swab positive | 6 (3.2) | 3 (2.0) |
| Saliva and SN swab positive | 4 (2.1) | 4 (2.7) |
| All three swabs positive in-tandem | 82 (43.6) | 15 (10.1) |
| Concordance of results among NP swabs (CDC-LDT), SN swabs (Fortitude 2.1), and saliva (Fortitude 2.1), *n* (%): |  |  |
| NP swab only positive | 9 (4.8) | 5 (3.4) |
| Saliva only positive | 14 (7.5) | 23 (15.4) |
| SN swab only positive | 2 (1.1) | 4 (2.7) |
| NP swab and saliva positive | 21 (11.2) | 6 (4.0) |
| NP and SN swab positive | 8 (4.3) | 2 (1.3) |
| Saliva and SN swab positive | 4 (2.1) | 1 (0.7) |
| All three swabs positive in-tandem | 84 (44.7) | 14 (9.4) |

NP; nasopharyngeal, SN; self-administered nasal, CDC-LDT; Centers for Disease Control and Prevention-Laboratory Developed Test

**Figure 1.**
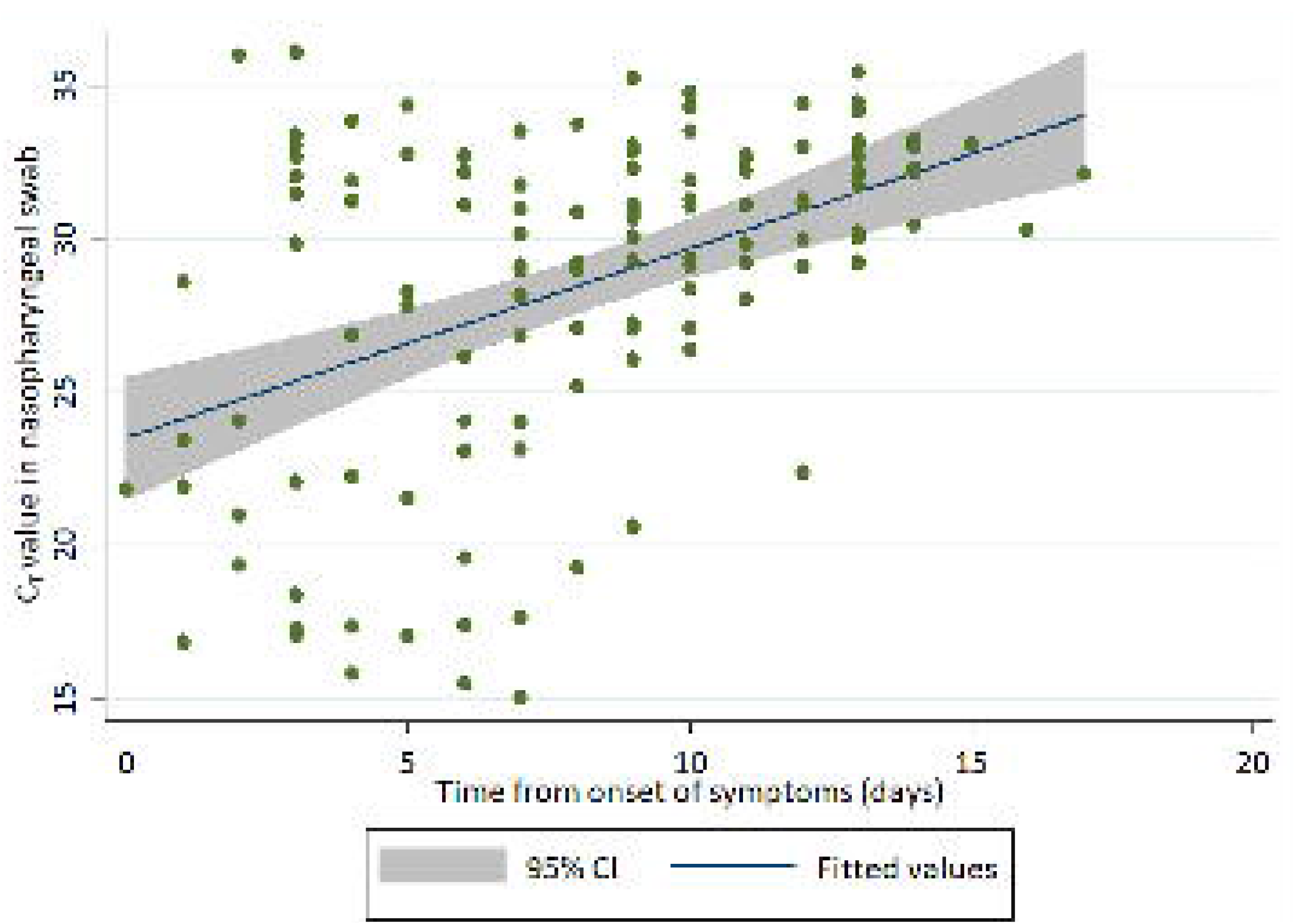
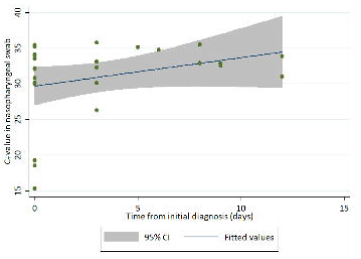
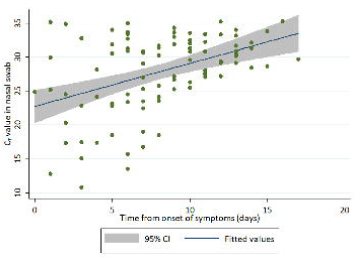
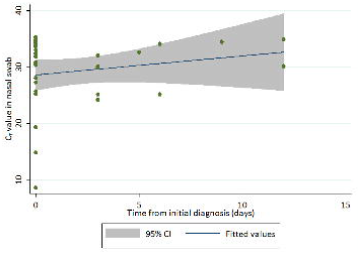
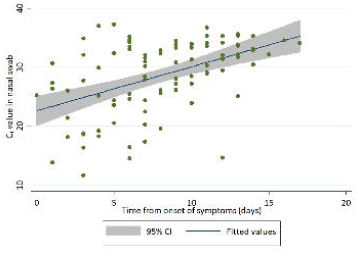
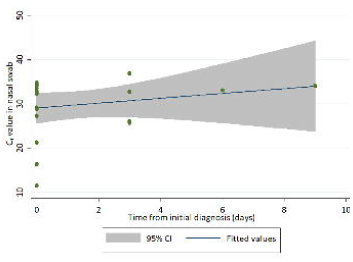
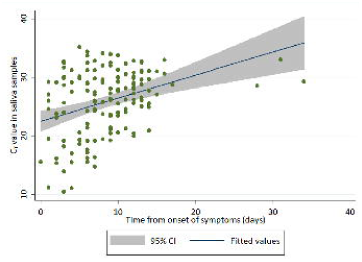
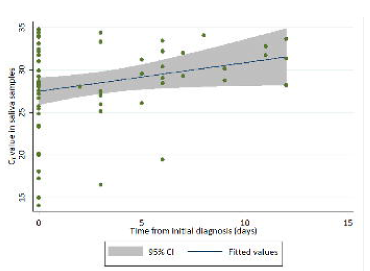
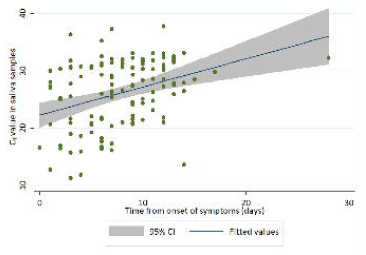
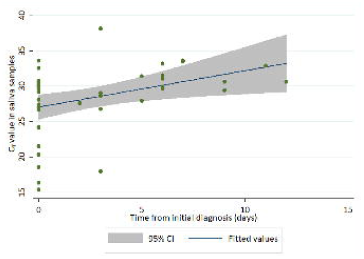
RT-PCR Ct values from onset of symptoms (symptomatic subjects) or time of diagnosis (asymptomatic subjects) for. A) NP swabs (tested via CDC-LDT) in symptomatic subjects B) NP swabs (tested via CDC-LDT) in asymptomatic subjects C) Self-administered nasal swabs (tested via CDC-LDT) for symptomatic subjects D) Self-administered nasal swabs (tested via CDC-LDT) from asymptomatic subjects E) Self-administered nasal swabs (tested via Fortitude 2.1) from symptomatic subjects F) Self-administered nasal swabs (tested via Fortitude 2.1) from asymptomatic subjects G) Saliva (tested via CDC-LDT) obtained from symptomatic subjects H) Saliva (tested via CDC-LDT) obtained from asymptomatic subjects I) Saliva (tested via Fortitude 2.1) obtained from symptomatic subjects J) Saliva (tested via Fortitude 2.1) obtained from asymptomatic subjects

For 63 positive NP swabs with low Ct (<30) values (the lower of results from two targets), SN swabs were concomitantly positive in 57 (90.5%) and 60 (95.2%) samples, while saliva was positive in 62 (98.4%) and 61 (96.8%) samples when tested via CDC-LDT and Fortitude 2.1, respectively. For 87 positive NP swabs with high Ct (≥30) values, SN swabs were concomitantly positive in 49 (56.3%) and 48 (55.2%) samples, while saliva was positive in 77 (88.5%) and 64 (73.6%) samples, respectively. The likelihood of concordant positive tests in other samples was also high when Ct values were low (<30) in saliva tested via CDC-LDT—76.9% of NP swabs, 64.0% of SN swabs tested via CDC-LDT, and 62.6% of SN swabs tested via Fortitude 2.1 were concomitantly positive. However, only 41.9% of SN swabs, 30.7% of SN swabs tested via CDC-LDT, and 27.4% of SN swabs tested via Fortitude 2.1 were positive in-tandem with saliva samples tested via CDC-LDT when Ct values were high (>30).

Test concordance between different sample sites was relatively good as evaluated by Cohen’s kappa coefficient, with the kappa statistic of 0.616 for NP and SN swabs tested via CDC-LDT (agreement of 81.3%), 0.675 for NP tested via CDC-LDT and SN swabs tested via

Fortitude 2.1 (agreement of 84.3%), 0.537 for NP and saliva tested via CDC-LDT (76.2%), and 0.602 for NP tested via CDC-LDT and saliva tested via Fortitude 2.1 (80.1%). Test concordances were excellent between the two saliva (87.2%) and SN swabs (91.0%) tests, with a kappa statistic of 0.745 and 0.806, respectively.

In the subset of confirmed COVID-19 subjects from the CCF, the likelihood of a positive test from any sample fell beyond 14 days of symptom onset in symptomatic subjects, although this was less significant for saliva tested via CDC-LDT (Table 3).

**Table 3.**
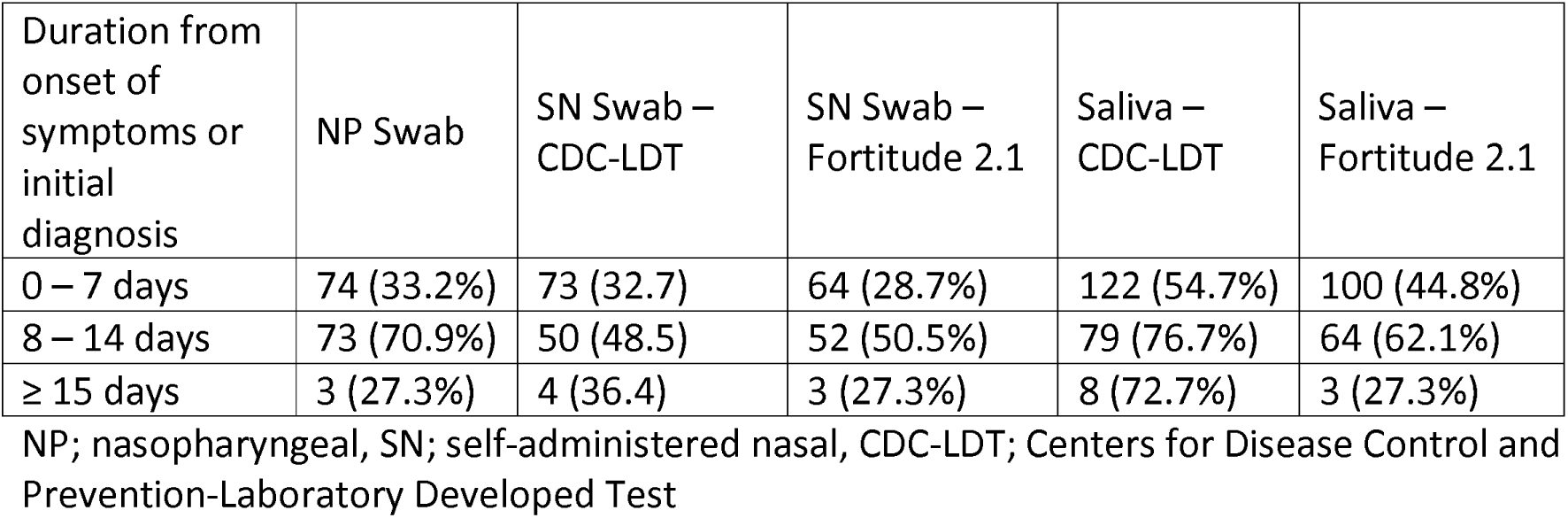
Likelihood of test positivity over time in confirmed COVID-19 subjects.

| Duration from onset of symptoms or initial diagnosis | NP Swab | SN Swab – CDC-LDT | SN Swab – Fortitude 2.1 | Saliva – CDC-LDT | Saliva – Fortitude 2.1 |
| --- | --- | --- | --- | --- | --- |
| 0 – 7 days | 74 (33.2%) | 73 (32.7) | 64 (28.7%) | 122 (54.7%) | 100 (44.8%) |
| 8 – 14 days | 73 (70.9%) | 50 (48.5) | 52 (50.5%) | 79 (76.7%) | 64 (62.1%) |
| ≥ 15 days | 3 (27.3%) | 4 (36.4) | 3 (27.3%) | 8 (72.7%) | 3 (27.3%) |

NP; nasopharyngeal, SN; self-administered nasal, CDC-LDT; Centers for Disease Control and Prevention-Laboratory Developed Test

Among 18 samples (SN swabs and saliva) from round 1 of sampling tested by NGS as proof of principle, there was a strong correlation between viral genome coverage (%) by NGS and Ct values for SARS-CoV-2, with 10 samples showing 100% coverage (7 unique subjects). Formal limit-of-detection of studies are pending.

Phylogenetic analyses of sequences of SARS-CoV-2 viral RNAfrom high-coverage saliva samples of unique subjects (n=7) was done, and the lineages for all the samples tested were determined to be Clade O by GISAID nomenclature [19] and lineage B.6 by PANGOLIN system of nomenclature [20].

## Discussion

Our study is concordant with multiple published works supporting saliva as an alternative sample for COVID-19 screening and diagnosis [7–15], and one of a minority where saliva was shown to be more sensitive than the corresponding NP swab [8,9,13], although the results by Leung and co-workers (53.7% saliva vs 47.4% NP swab, 95 subjects) were not statistically different [8].

Several reasons may account for this difference in these studies, including the strict collection of saliva that includes oropharyngeal secretions where the viral load is potentially higher [8,9], or a higher volume of collection in the Yale study, where 1/3 of a urine cup (approximately 10 mls) had to be filled with saliva [13]. Because NP swabs were performed by trained healthcare staff locally, it is less likely that poor NP swab technique resulted in lower yield. The laboratory performing saliva testing similarly performed testing for all SN and most of the N P swabs for this study, and an extensive review including environmental testing revealed no evidence of laboratory contamination. Dithiothreitol (DTT) was used to pre-process all saliva samples here before RNA extraction, to resolve the commonly reported issue of saliva specimen viscosity, which can lead to false-negatives.

Interestingly but perhaps unsurprisingly, the use of different RT-PCR kits resulted in different test-positive rates in saliva in our study, suggesting that this can potentially be an important consideration for clinical laboratories, where more sensitive laboratory protocols should be deployed for clinical diagnosis as opposed to mass screening for low-prevalence populations. More validation will be required to confirm this.

SN swabs, however, appeared less sensitive compared to both NP swabs and saliva for the diagnosis of COVID-19. Although this was convenient, less time-consuming to perform relative to saliva collection and caused less discomfort compared to NP swabs, the markedly lower sensitivity should preclude its use where other sample types can be collected.

In our study, NGS can provide whole-genome profiling of -CoV-2 directly from clinical samples for phylogenic analysis. Formal limit-of-detection studies are still pending for the NGS workflow here; other groups have reported highly sensitive performance for NGS where a threshold of 5% genome coverage or 84 genome-equivalents per ml., was established as limit of detection [21]. The phylogeny results were consistent with the virus belonging to viral type (Clade 0, lineage B.6), known to be circulating in the geographical regions of Singapore and India.

There are several limitations to our work. Firstly, the study population was confined to young and middle-aged men who were either asymptomatic or had mild disease. The results cannot be extrapolated to other populations where there is a clear need for alternate sample types to NP swabs, such as in the paediatric population. Secondly, we did not extend the follow-up testing sufficiently to determine when saliva viral shedding stopped for the majority of subjects, although this has been explored in other studies [7,10]. Thirdly, we did not test for the difference, if any, between naso-oropharyngeal saliva collection and saliva obtained from the mouth alone, although it is biologically plausible that the latter would result in lower sensitivity for COVID-19 diagnosis.

In conclusion, our study adds to the considerable body of evidence supporting saliva as a sensitive and less intrusive sample for COVID-19 diagnosis and further defines the role of oropharyngeal secretions and impact of different RT-PCR kits in increasing the sensitivity of testing. In our study, SN swabs were inferior as an alternate sample type to both NP swabs and saliva. Our study also provides evidence that a straightforward next-generation sequencing workflow can provide direct-from-sample phylogenetic analysis for public health decision-making.

## Data Availability

All test data in the manuscript are available for sharing on request.

## Acknowledgments

We would like to thank S Siva Ranjini and Lenny Indrawati of Saw Swee Hock School of Public Health, the Singapore General Hospital, Woodlands Health Campus, and Singapore Armed Forces Medical Corp for supporting and facilitating the work at the purpose-built dormitory and CCF. We are also very grateful for the support and patience of the migrant workers who participated in this study, and acknowledge the uncertainty and difficulties they have experienced as a consequence of COVID-19.

